# Effects of Home-Based EEG Neurofeedback Training as a Non-Pharmacological Intervention for Parkinson’s Disease

**DOI:** 10.1101/2024.02.19.24303031

**Authors:** Andrew Cooke, John Hindle, Catherine Lawrence, Eduardo Bellomo, Aaron W. Pritchard, Catherine A. MacLeod, Pam Martin-Forbes, Sally Jones, Martyn Bracewell, David E. J. Linden, David M. A. Mehler

## Abstract

Aberrant movement-related cortical activity has been linked to impaired motor function in Parkinson’s disease (PD). Dopaminergic drug treatment can restore these, but dosages and long-term treatment are limited by adverse side-effects. This experiment reports the first study of home-based electroencephalographic (EEG) neurofeedback training as a non-pharmacological candidate treatment for PD. Sixteen people with PD received six home visits comprising symptomology self-reports, a standardised motor assessment, and a precision handgrip force production task while EEG was recorded (visits 1, 2 and 6); and 3 × 1-hr EEG neurofeedback training sessions to reduce EEG high-alpha power before initiating handgrip movements (visits 3 to 5). Participants successfully learned to self-regulate movement-related alpha rhythms, and this appeared to expedite the initiation of precision movements. There was no evidence of wider symptomology reduction. Interviews indicated that the intervention was well-received. We conclude that home-based neurofeedback for people with PD is feasible and warrants further research.

## Introduction

Idiopathic Parkinson’s disease (PD) is a neurodegenerative condition that impairs motor function. PD is the second most common cause of chronic neurological disability after stroke, affecting 1.5% of people aged over 70 years^1^. Its main motor symptoms include akinesia (loss or impairment of voluntary movement), bradykinesia (fatigable slowness of movement), resting tremor, rigidity, and loss of postural reflexes^2^. The pathophysiology of PD is characterised by the progressive decay of dopamine-producing neurons in the substantia nigra region of the brain^3^. Dopaminergic medication (e.g., levodopa and dopamine agonists) is typically deployed to alleviate some of the motor symptoms of PD^4^. However, people with PD can show variations in response and prolonged pharmacological treatment is often associated with motor (e.g., involuntary movements known as dyskinesias) and psychiatric (e.g., confusion, hallucinations, apathy and somnolence) side-effects^5^. For instance, up to 40% of people with PD may be affected by dyskinesia after 4 to 6 years of levodopa (L-Dopa) treatment^6^. The combination of drugs and disease progression may trigger apathy and somnolence in up to 20% and 50% of people with PD, respectively^7,8^. Moreover, pharmacotherapy dosages must be increased over time to maintain effectiveness^9^, which may further increase the risk for developing side effects. These dose-limiting effects underscore a pressing need for the development of sustainable, non-pharmacological approaches, such as neuromodulation techniques, to manage motor symptoms of PD. However, studies of effective non-pharmacological treatments are very limited^10^.

Non-invasive neurofeedback training in PD is a promising, personalised, non-pharmacological intervention that aims to enable individuals to modulate their brain activity^11^. With a lower likelihood of side-effects, when compared to pharmacological treatments, neurofeedback training is a possible alternative or adjunct to medication, with the potential to improve the quality of life for people with PD^12^. Initial studies of motor imagery based real-time functional magnetic resonance imaging (rt-fMRI) neurofeedback have shown promising results in enabling people with PD to increase the activation of the supplementary motor area (SMA)^13,14^, a cortical brain region that shows reduced activation in people with PD during preparation and execution of voluntary movements^15,16^. Further, SMA based rt-fMRI neurofeedback training was associated with significant reductions in PD motor symptoms^13,14^. However, limitations of this approach include the need for access to an MRI scanner, its high costs, as well as contraindications (e.g., exclusion of people who have ferromagnetic implants). In contrast, scalp-based electroencephalography (EEG) is cheaper, has essentially no contraindications and is widely available in neurological health care settings. Crucially, the portability of EEG presents opportunities for decentralised clinical trials such as home-based EEG neurofeedback treatment, which may aid recruitment and retention^17^. Moreover, decades of observational and intervention studies that employed EEG recordings in people with PD provide observational and mechanistic insights that can inform the choice of a suitable EEG neurofeedback target signal^18^.

One particularly promising candidate signal for EEG neurofeedback training in PD is alpha event-related desynchronization (ERD). ERD describes the notable reduction in power in the alpha band (around 8-12 Hz) at central electrode locations overlying the primary motor areas that immediately precedes voluntary movement^18^. People with PD show attenuated alpha ERD at contralateral sensorimotor areas (i.e., C3 or C4) compared to healthy controls^19^. Importantly, dopaminergic medication helps to restore alpha ERD amplitudes^16,20^ and alpha ERD correlates with improved bradykinesia symptoms^21^. Further, deep brain stimulation studies that targeted the subthalamic nucleus in combination with L-Dopa have shown an increase in alpha ERD latency during movement preparation and an increase in alpha ERD during movement execution over central electrodes^22^. Taken together, these findings suggest that: a) pre-movement alpha ERD, particularly at electrode sites overlying contralateral sensorimotor areas (e.g., C3 electrode for right hand movements, C4 electrode for left hand movements), is a key determinant of the preparation and initiation of movement; b) PD disrupts “normal” pre-movement alpha ERD, and this could explain movement difficulties; and c) current pharmacological therapy can restore “normal” pre-movement alpha ERD and motor performance. Pre-movement alpha ERD over the motor cortex is thus considered a promising EEG marker to target motor impairment in PD.

Importantly, research in healthy controls has established that pre-movement alpha ERD is trainable via relatively brief neurofeedback interventions. For example, healthy participants were able to exert some control over their pre-movement alpha ERD after just 30-mins of central alpha neurofeedback training^23^. This was associated with an immediate benefit in cognitive (serial-sevens accuracy) and motor (timed-up-and-go test) performance^23^. Another study provided 3-hrs of frontocentral alpha ERD training to healthy participants and found a strong and enduring training effect where participants were able to reduce their pre-movement frontocentral alpha for at least a few days after the feedback was withdrawn^24^. Surprisingly, however, only a handful of exclusively laboratory-based studies attempted scalp-based EEG neurofeedback for PD, as described in detail in recent reviews^12,18,25^. Although these preliminary studies provide some initial evidence for the feasibility of EEG neurofeedback training in people with PD in a laboratory setting, they have been largely inconclusive due to insufficient clinical information (e.g., assessing/reporting symptom changes), very small sample sizes (mostly case studies), only single training sessions, and lack of inferential statistical testing^25^. Crucially, the choice of neurofeedback target varied between (and in some cases, within) previous studies, with none of them targeting central alpha ERD despite the compelling case for this neurofeedback target. Given the initial feasibility of alpha ERD neurofeedback training in healthy participants and the therapeutic potential of alpha ERD neurofeedback training for PD, we aimed to test the feasibility of alpha-based EEG neurofeedback training in people with PD in the present study.

We took several steps to provide a highly original and rigorous examination of EEG neurofeedback in PD. First, based on a thorough review of previous literature, we identified alpha ERD as an evidence-based target for training^18^. Second, our design was informed by studies that successfully trained alpha ERD in healthy participants^23,24^. Third, the details of our study were pre-registered (https://www.isrctn.com/ISRCTN16783092). Fourth, we evaluated our protocol in accord with the recent consensus on the reporting and experimental design of clinical and cognitive-behavioural neurofeedback studies (CRED-nf) checklist (see supplementary material), to help promote robust experimental design and transparent reporting^26^. Finally, and most importantly, the present experiment was designed to deliver the neurofeedback intervention in participants’ homes, making it the first examination of portable, home-based neurofeedback treatment in PD.

This study provides a multi-session design comprising test and training phases in people with PD. Their motor performance, PD symptomology and EEG activity was assessed in their homes while they were taking their normal anti-Parkinsonian medicine (pre-test A), and while they were off their medication during a visit before (pre-test B) and after (post-test) a course of neurofeedback training. In-between pre-test A and post-test, participants completed three separate 1- hr sessions of alpha ERD neurofeedback training (i.e., training phase) while we assessed their cortical activity and their motor performance. Specifically, following successful self-regulation, participants performed a precision handgrip motor task. Hence, this novel design combined classical neurofeedback training with a relevant motor execution task. Overall, the study comprised six home visits per participant, each separated by at least 2 days, and was designed to afford some insight into the relative effects of anti-Parkinsonian medication (pre-test A versus pre-test B) and EEG neurofeedback training (pre-test B versus post-test).

The primary aim of the study was to test the feasibility of our neurofeedback intervention in a home setting. We hypothesized quadratic effects in the test phase of the design, where EEG alpha power would be greater, indicative of impaired ERD, during (off-medication) pre-test B compared to (on-medication) pre-test A and that this difference would normalise to some degree during the (off-medication) post-test following the neurofeedback training. This pattern would provide novel evidence of learned self-regulation via home-based training. Motor performance and symptomology were our secondary outcomes. We explored whether these were characterised by similar levels in pre-test A and post-test, and worse motor performance and more severe symptomology in pre-test B. Such findings would evidence an ability of alpha ERD neurofeedback training to yield similar outcomes to PD medication. We further hypothesised linear effects in the training phase, characterised by a decrease in alpha power and an improvement in motor performance from one training session to the next. Finally, as this is the first study of EEG neurofeedback in participant homes, we also assessed the qualitative experience of participants via a brief interview.

## Results

### Part A: Quantitative

#### Cortical Activity

##### Test Phase

Repeated measures Polynomial Trend ANOVAs performed for each recording electrode revealed significant quadratic effects at the C3 (*F*(1,12) = 5.87, *p* =.03, η_p_^2^ = .33) and the C4 (*F*(1,12) = 5.50, *p* =.04, η_p_^2^ = .31) sites, and directionally similar non-significant effects at Cz (*F*(1,12) = 4.02, *p* = .07, η_p_^2^ = 27) and Fz (*F*(1,12) = 1.09, *p* = .32, η_p_^2^). All sites were characterised by an increase in high-alpha power from pre-test A to pre-test B, and then a decrease in high-alpha power from pre-test B to post-test (Figs. 1A - 1D). This provides evidence that the neurofeedback intervention was successful in shaping participants’ EEG high-alpha power in the prescribed manner at the targeted bilateral C3 and C4 sites.

**Figure 1.**
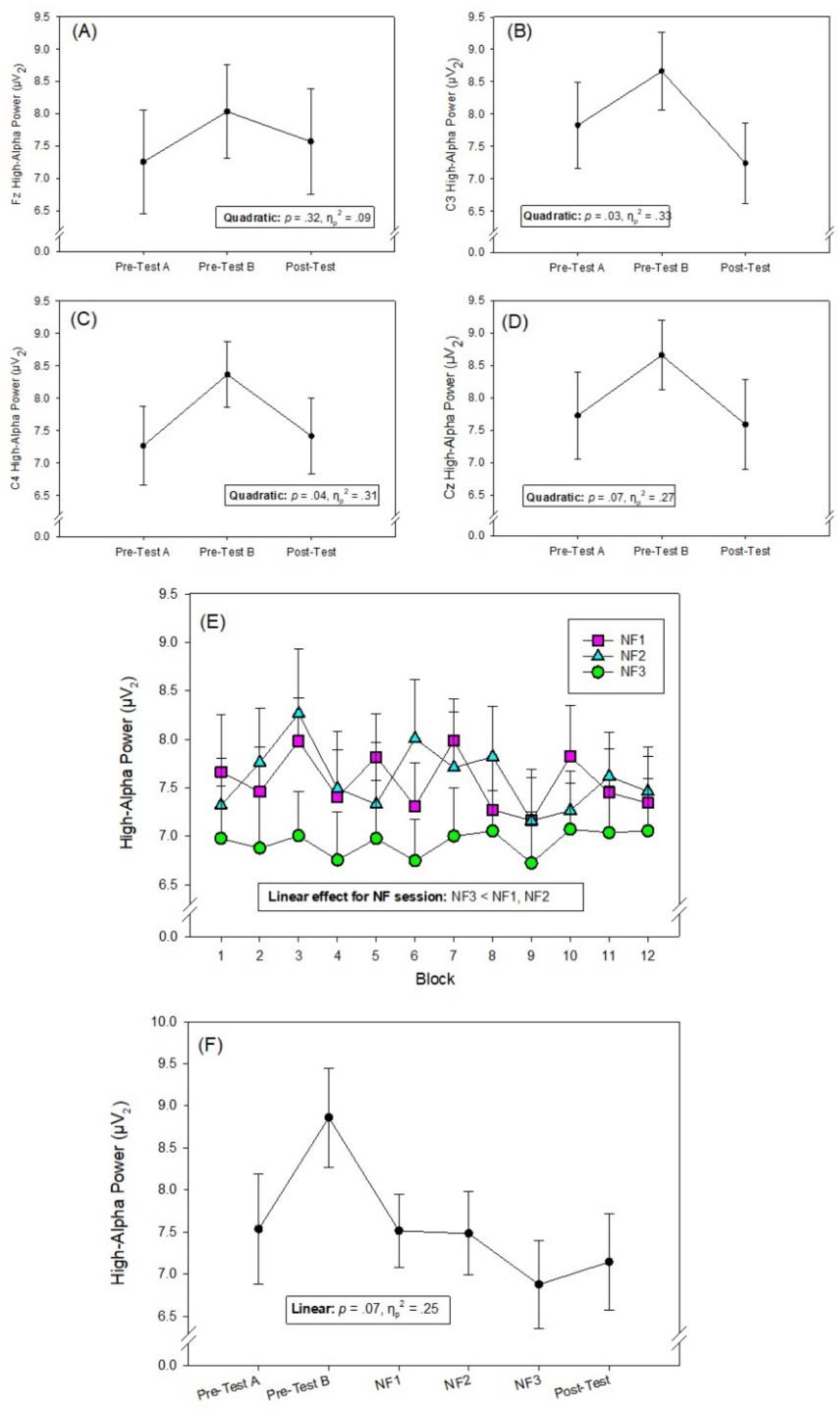
Mean EEG high-alpha power recorded during the test phase of the experiment for different electrode positions (Panels A-D), the training phases of the experiment for EEG neurofeedback electrode position C3 (C4 in left handers – Panel E), and across all experimental visits for EEG neurofeedback electrode position C3 (C4 in left handers – Panel F). Pre-test A = on anti-Parkinsonian medication and before neurofeedback intervention. Pre-test B = off anti-Parkinsonian medication and before neurofeedback intervention. NF1, NF2 and NF3 = neurofeedback training sessions off anti-Parkinsonian medication. Post-test = off anti-Parkinsonian medication and after neurofeedback intervention. Error bars indicate standard error of the means.

##### Training Phase

The 3 × 12 Polynomial Trend ANOVA employed to analyse cortical activity during the neurofeedback training phase of the experiment revealed a significant linear effect for Session (*F*(1,14) = 4.97, *p* =.04, η_p_^2^ = .26), showing a decrease in C3 (C4 for left-handers) high-alpha power from the first and second neurofeedback sessions to the third neurofeedback session (*M*_session 1_ = 7.56 µV^2^, *M*_session 2_ = 7.60 µV^2^, *M*_session 3_ = 6.94 µV^2^ per block). This provides evidence for a between-session effect of the neurofeedback intervention; participants were most effective in suppressing their high-alpha power in the final training session where the feedback threshold was the most severe (Fig. 1E). There was no significant linear effect for Block (*F*(1,14) = 0.19, *p* =.67, η_p_^2^ = .01) and there was no Session × Block interaction, (*F*(1,14) = 1.17, *p* =.30, η_p_^2^ = .08). These data imply that our neurofeedback intervention was successful in encouraging self-regulation of high-alpha power, with adaptations occurring between, but not within, individual training sessions. Additional analyses based on the number of silenced tones revealed linear effects for Session and Block (see supplementary material).

##### All Visits

A six-level repeated measures ANOVA to compare high-alpha power at the neurofeedback site over the six visits that comprised both the test and the training phase of the experiment revealed a significant effect of visit (*F*(5,60) = 3.04, *p* =.04, η_p_^2^ = .20, ε = .56). This was characterized by a marginal linear trend (*F*(1,12) = 4.00, *p* = .07, η_p_^2^ = .25), where high-alpha power tended to be greatest in pre-test B, and lowest in the final neurofeedback training session and the post-test (Fig. 1F).

#### Precision Handgrip Performance

##### Test Phase

Repeated measures Polynomial Trend ANOVAs did not reveal the hypothesised quadratic effects: movement planning time (*F*(1,14) = 0.39, *p* =.55, η_p_^2^ = .03); absolute error (*F*(1,14) = 2.63, *p* =.13, η_p_^2^ = .16); constant error (*F*(1,14) = 0.99, *p* =.34, η_p_^2^ = .07); variable error (*F*(1,14) = 1.23, *p* =.28, η_p_^2^ = .08). There were, however, significant linear effects for movement planning time (*F*(1,14) = 4.88, *p* =.04, η_p_^2^ = .26) and for variable error (*F*(1,14) = 5.26, *p* =.04, η_p_^2^ = .27). Inspection of the means revealed that participants were able to produce grip forces typically within 1% MVC of their target, but with a slight bias to under-squeeze the handgrip dynamometer. Importantly, their movement planning times reduced, and they became more consistent from the pre-tests to the post-test, providing evidence of improved movement planning and initiation across the Test phase (Figs. 2A - 2D).

**Figure 2.**
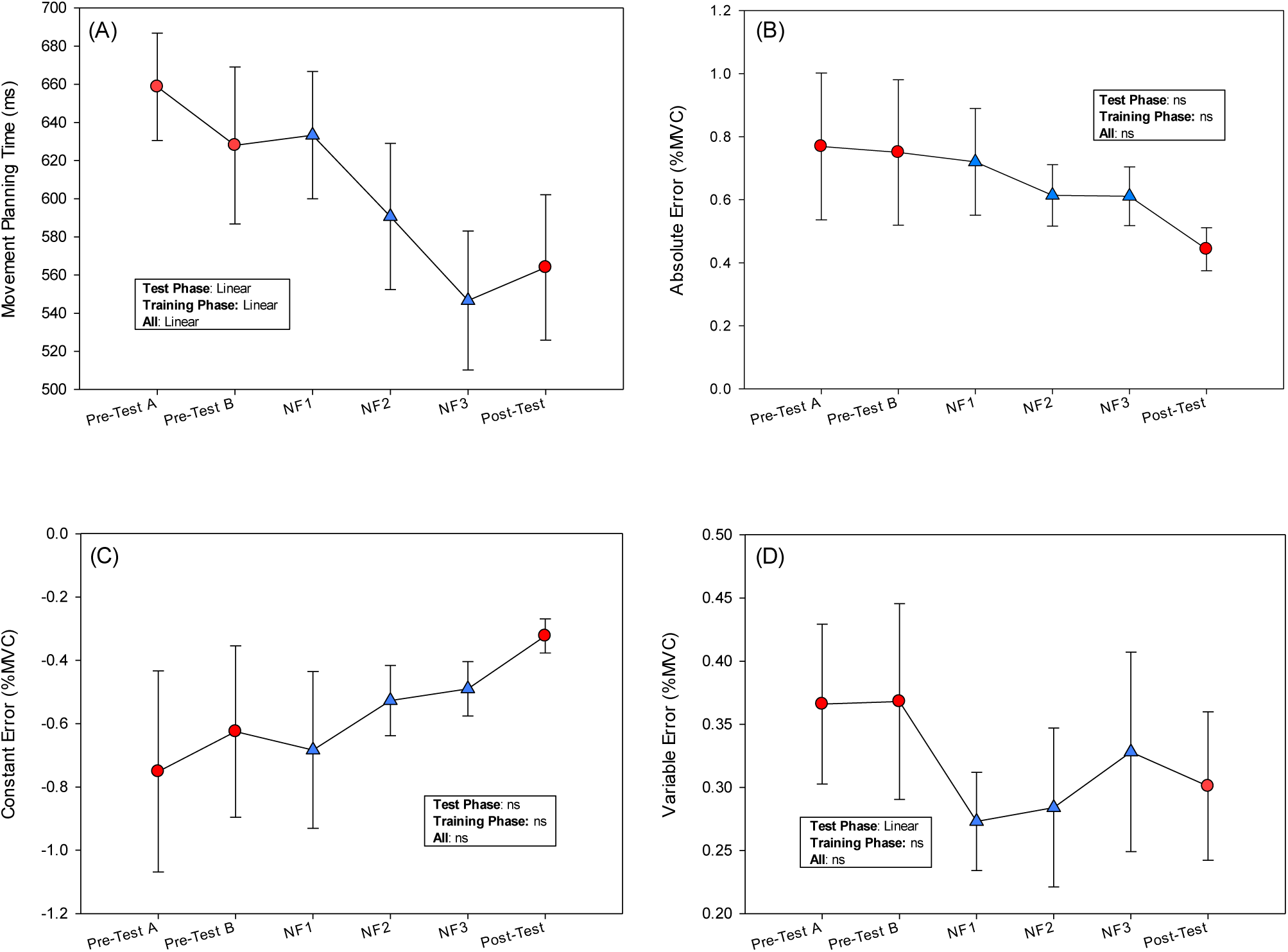
Mean movement planning time (Panel A), absolute error (Panel B), constant error (Panel C), variable error (Panel D) and on the precision handgrip task. For movement planning time, smaller values indicate better performance. Absolute error reflects accuracy (i.e., proximity to target), constant error reflects bias (i.e., any tendency to undersqueeze or oversqueeze) and variable error reflects consistency – in all cases scores of zero (no error, no bias, strong consistency) is optimal. Pre-test A = on anti-Parkinsonian medication and before neurofeedback intervention. Pre-test B = off anti-Parkinsonian medication and before neurofeedback intervention. NF1, NF2 and NF3 = neurofeedback training sessions off anti-Parkinsonian medication. Post-test = off anti-Parkinsonian medication and after neurofeedback intervention. Error bars indicate standard error of the means. *Note*: Red circles indicate three data points analysed in the test phase. Blue triangles indicate three data points analysed in the training phase. All six datapoints were analysed in the “All” phase.

##### Training Phase

Repeated measures Polynomial Trend ANOVAs revealed the expected significant linear effect for movement planning time (*F*(1,14) = 14.32, *p* =.002, η_p_^2^ = .51). This was characterized by an improvement in performance (i.e., shorter planning times) from neurofeedback training session 1 to training session 2, and again from training session 2 to training session 3 (Fig. 2A). There were no linear effects for absolute error (*F*(1,14) = 0.84, *p* =.37, η_p_^2^ = .06), constant error (*F*(1,14) = 1.12, *p* =.31, η_p_^2^ = .07) or variable error (*F*(1,14) = 1.47, *p* =.25, η_p_^2^ = .10).

##### All Visits

Six-level repeated measures ANOVAs to compare performances across all experimental visits revealed a significant effect for movement planning time (*F*(5,70) = 4.40, *p* =.009, η_p_^2^ = .24, ε = .61). This was characterized by a significant linear trend (*F*(1,14) = 9.04, *p* =.009, η_p_^2^ = .39), with improvements in performance over the course of the experiment, and which were particularly evident during the training phase (Fig. 2A). There were no significant effects for absolute error (*F*(5,70) = 1.87, *p* =.19, η_p_^2^ = .12, ε = .26), constant error (*F*(5,70) = 1.65, *p* =.22, η_p_^2^ = .11, ε = .24) or variable error (*F*(5,70) = 2.77, *p* =.07, η_p_^2^ = .16, ε = .49).

##### MDS-UPDRS and PDQ-8

The mean scores for MDS-UPDRS Part II (motor aspects of experiences of daily living), the MDS-UPDRS Part III (motor examination) and the PDQ-8 are presented in Table 1. Polynomial trend ANOVAs revealed significant quadratic and linear trends for the MDS-UPDRS Part III, (Quadratic: *F*(1,14) = 5.36, *p* =.04, η_p_^2^ = .28; Linear: *F*(1,14) = 8.28, *p* =.01, η_p_^2^ = .37) with the effect sizes indicating that the linear trend was strongest. This effect was driven by the least severe symptomatology observed in the (on-medication) pre-test A, and most severe symptomology observed in the (off-medication) post-test (Table 1). No significant trends emerged for the self-reported measures, MDS-UPDRS Part II (Quadratic: *F*(1,14) = 0.71, *p* =.41, η_p_^2^ = .05; Linear: *F*(1,14) = 1.70, *p* =.21, η_p_^2^ = .11), PDQ-8, (Quadratic: *F*(1,14) = 1.09, *p* =.32, η_p_^2^ = .07; Linear: *F*(1,14) = 1.11, *p* =.31, η_p_^2^ = .07).

**Table 1.** MDS-UPDRS and PDQ-8 Descriptive Statistics.

| Measure (range of possible scores)<sup>a</sup> | Pre-test A<br>(on-medication) |  | Pre-test B<br>(off-medication) |  | Post-test<br>(off-medication) |  |
| --- | --- | --- | --- | --- | --- | --- |
|  | *M* | *SD* | *M* | *SD* | *M* | *SD* |
| MDS-UPDRS Part II (MEDL) (0-52) | 10.94 | 8.79 | 9.63 | 7.37 | 9.53 | 7.10 |
| MDS-UPDRS Part III (motor examination) (0-132) | 22.88 | 14.56 | 29.00 | 14.75 | 30.07 | 11.99 |
| PDQ-8 (0-100) | 18.36 | 15.13 | 20.12 | 17.04 | 17.50 | 13.92 |

*Note:* <sup>a</sup>Higher scores indicate more severe motor symptoms of PD (MDS-UPDRS) / reduced quality of life (PDQ-8).

**Table 2.** Participant Inclusion and Exclusion Criteria.

| Inclusion Criteria | Exclusion Criteria |
| --- | --- |
| 1. Idiopathic PD diagnosed using the UK Brain Bank Diagnostic Criteria<br>2. Hoehn and Yahr stage 1-3<br>3. On stable treatment for the past month and no planned changes over the period of the study<br>4. Ability to give informed consent<br>5. Aged over 18 years | 1. Clinical diagnosis of dementia<br>2. Active psychosis, clinically significant depression or behavioural disturbance<br>3. Presence of another neurological condition |

### Part B: Qualitative

In response to the questions posed at the end of the neurofeedback training, 8 out of 15 participants (56%) felt that they perceived some benefits of the neurofeedback training, while 13 out of 15 participants (86%) indicated that they would recommend this type of neurofeedback training to other people with PD. Thematic content analyses identified three areas of perceived benefit: improved walking, improved psychological control, and improved motor control. These themes and some example comments related to each are presented in Fig. 3.

**Figure 3.**
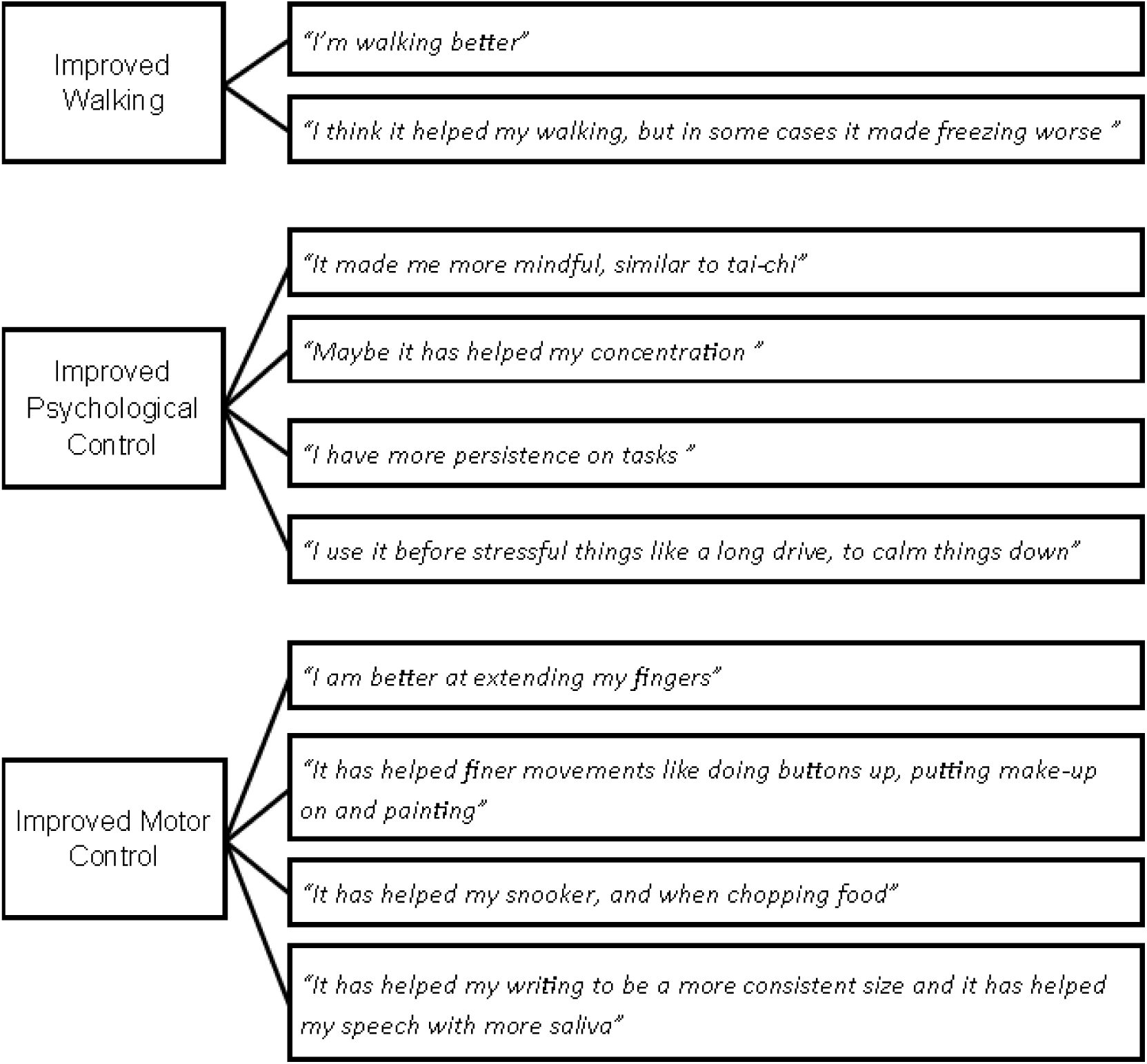
Perceived benefits of neurofeedback training and example quotes from participants.

Reasons that participants offered to support their assertion that they would recommend this type of neurofeedback training to others included: *“it is good to find non-pharmacological treatments, it empowers you”*, *“it is fun”*, *“having visual and auditory feedback is highly useful”*, *“it is interesting”*, *“controlling the alpha makes you able to control the subconscious”* and finally, *“because it works for me!”*.

The final question, probing neurofeedback strategies, was explorative. In brief, 12 out of 15 participants (80%) reported developing some helpful strategies to silence the neurofeedback tone (i.e., control their alpha ERD). The most reported strategy was a combination of focus on the neurofeedback screen along with strategies to relax and clear the mind of anything else. These explorative data are reported in Supplementary Table 1.

## Discussion

The primary goal of this experiment was to provide the first examination of the feasibility of home-based EEG alpha ERD-neurofeedback training in people with PD. It represents a unique decentralised trial of a scalable, non-invasive, and personalised neuromodulation technique. Our target for neurofeedback training, ERD in the alpha frequency band, was informed by previous research implicating pre-movement ERD at central sites as key indices of movement efficacy in people with PD (e.g.,^16,18,19^). Based on earlier reports from healthy controls, we hypothesised that central EEG alpha power would be trainable^23^ in a home setting. Our secondary aims were to shed light on the potential efficacy of neurofeedback as a candidate non-pharmacological treatment for the motor symptoms of PD (e.g., explore the comparative effects of neurofeedback versus pharmacological treatment on PD symptomology). Finally, we used thematic content analyses to qualitatively explore participants’ experience of this novel home-based EEG neurofeedback training.

### Effects of Neurofeedback on Cortical Activity

Based on our hypotheses, we expected that quadratic changes (test-phase) and linear changes (training-phase) in central alpha power would evidence learned self-regulation and endorse the feasibility of our neurofeedback treatment. EEG data during the test phase of the study confirmed the expected quadratic effect where high-alpha power was greater (suggesting impaired ERD) in pre-test B (off-medication) than in pre-test A (on-medication) and post-test (off-medication) at the bilateral C3 and C4 sites. The difference between the pre-tests is in line with previous research that reported improved alpha ERD before and during finger and hand movements when participants were on anti-Parkinsonian medication^16,20^. Importantly, our home-based EEG neurofeedback training appeared to enable people with PD to reduce their pre-movement high-alpha power in the absence of medication (post-test). This finding is in line with previous work that demonstrated the trainability of EEG alpha power in healthy controls^23,24^ and it provides encouraging evidence that EEG alpha neurofeedback training can help people with PD to self-regulate their pre-movement cortical activity over the C3 (or C4) electrode position.

Data obtained during the training phase of the experiment provide further support for the efficacy of the intervention in training participants to self-regulate their cortical activity. Specifically, there was a linear effect for training session whereby high-alpha power decreased (suggestive of greater ERD) from the first and second to the final neurofeedback training session. This could reflect some self-regulation of EEG activity generated by the primary motor cortex^27^, a key region involved in encoding kinematic and muscle information of complex hand movements^28^. Previous attempts of motor imagery-based rt-fMRI neurofeedback training that targeted the primary motor cortex yielded effects that were mostly restricted to premotor areas^13,14^, suggesting neurophysiological differences between imagery-based and pre-movement neurofeedback approaches. High-alpha power did not change significantly within each training session, although there was some evidence for within-session improvement indexed by more frequent silencing of the tone as each session progressed (supplementary Fig. 1). It is likely that sleep consolidation accelerated training benefits^29^, which may explain why the between-session effects were stronger than the within-session changes. To capitalize on this effect, future research could implement a higher number of shorter duration training sessions, spread over a greater number of days.

In sum, the experiment provides very encouraging EEG data to reveal for the first time that participants with PD can use home-based neurofeedback to restore disrupted pre-movement high-alpha rhythms to a degree that is comparable to the effects of pharmacological intervention (Fig. 1B). We hence achieved our first aim, and our results provide the first confirmation of the feasibility of home-based neurofeedback training in PD.

### Effects of Neurofeedback on Motor Performance and PD Symptomology

Our secondary outcomes concerned the effects of the neurofeedback treatment on measures of motor performance and PD symptomology. We expected our measures of precision handgrip performance to yield quadratic effects during the test phase of the experiment to index a worsening performance from pre-test A to pre-test B (reflecting the withdrawal of medication) and an improvement from pre-test B to post-test (reflecting benefits of neurofeedback training). Instead, we found linear improvements for movement planning time (all phases) and variable error (test phase), and no significant changes for absolute error and constant error. While not in line with our initial expectations, it is noteworthy that the clearest performance effects were the linear reductions in movement planning time. Reduced movement planning time reflects improved performance due to superior pre-planning of force^30,31^ and alpha ERD is closely related to planning and initiation of movement^32^. One could attribute this performance improvement simply to practice. However, the improvements occurred when participants were off-medication, which has previously stifled learning^33^. Moreover, if practice were the complete explanation for our movement planning time results, we would expect to see similar practice effects across the other performance measures. The inconsistent improvement in variable error (i.e., limited to the test phase – Fig. 2D) and the absence of improvement across the absolute and constant error measures thereby challenges the practice effect interpretation. Instead, our findings indicate a selective effect of our neurofeedback protocol on movement planning, where benefits accrued from one neurofeedback training session to the next, and where some of those benefits were retained at post-test when the neurofeedback was withdrawn (Fig. 2A).

In line with this interpretation of the data, it is worth noting that our observer-rated, rather global behavioural measure of PD symptomology (MDS-UPDRS Part III), was unaffected by our intervention (i.e., similar symptomology in pre-test B and post-test). This adds further credence to the idea that benefits of our neurofeedback protocol were very specific to the planning of our target handgrip task. Finally, neither medication nor neurofeedback training resulted in a significant change in self-report measures related to motor aspects of daily living or quality of life. These measures ask respondents to complete the items in relation to their feelings over the past week (MDS-UPDRS Part II) and the past month (PDQ-8). This method of aggregate recall over time may have washed out the relatively transient increases in symptomatology that participants were experiencing (i.e., as captured in the MDS-UPDRS Part III) during the off-medication visits.

### Participant Experience of Neurofeedback Training

In addition to the quantitative data, we obtained brief qualitative data and vignettes to investigate participants’ personal accounts and experience of our home-based intervention. These data indicate that the intervention was well-received. Just over half of the participants expressed that they perceived some benefit of the neurofeedback training, with the key themes being improved walking, improved psychological control (e.g., ability to concentrate and relax) and improved motor control (Fig. 3). In addition, nearly all participants (13/15) indicated that they would recommend EEG neurofeedback training to other people with PD. This suggests that most participants were pleased with the intervention, and this was backed up by comments indicating that the neurofeedback was “interesting”, “empowering” and “fun”. The lack of a control group makes it difficult to exclude the possibility of socially desirable responding that could have inflated the positive sentiment in the qualitative reports^26,34^. However, it is plausible that the home-based nature of the training did instil some genuine psychological benefits. For instance, in accord with the tenets of basic needs theory^35^, the regular social interaction with the experimenter may have helped participants to fulfil the basic psychological need for relatedness. Learning to self-regulate brainwaves may enhance perceived psychological control and assist in satisfying basic needs for autonomy and competence. Furthermore, any combination of these factors can inflate perceived coping resources, allowing individuals to appraise situations (e.g., how one feels right now, with current symptomology), in more favourable ways^36^. Recent neurofeedback studies support these theoretical assertions, with successful self-regulation itself (e.g., with a relaxation technique) being shown to yield improvements for different mood states^37^, which can occur with just a single training session^38^. This could explain why self-reported symptomology was similar at post-test (off-medication) compared to pre-test A (on-medication), even though observed symptomology at post-test was worse. In addition to the qualitative reports, it is also noteworthy that all but one participant completed the multi-session protocol, attesting to the potential for superior retention-rates of participants in decentralised compared to traditional clinical trials^17^. In sum, the views captured from our sample indicate that future home-based non-pharmacological interventions such as EEG neurofeedback are likely to be well received by people with PD.

### Future Directions

This feasibility study demonstrates the practicality and potential utility of home-based EEG neurofeedback training in PD and can provide a blueprint for future research to further interrogate EEG neurofeedback as a non-pharmacological PD treatment. We have already highlighted that future research would do well to replicate our experiment with a control group to better disentangle specific intervention benefits from those that may arise due to practice or socially desirable responding^25,31^. In addition, future neurofeedback research could consider providing training in augmentation to medication. For instance, honing self-regulation skills via neurofeedback while being on-medication may eventually allow a reduction in medication dosage, with the EEG entrainment supplementing the reduced dosage to yield equivalent motor outcomes. On the basis that medication can enhance learning^33^, an on-medication state may enhance neurofeedback training success. Neurofeedback training success may further benefit from providing explicit strategies to guide participants in how to control the neurofeedback signal from the start^39^. In reporting strategies that participants identified by themselves in the present study (supplementary Table 1), we provide some useful starting points for future work.

While the present intervention impacted most clearly upon movement initiation, alternative neurofeedback protocols that focus on EEG activity during movement execution may be able to yield benefits to online control measures. For example, protocols could provide continuous alpha power feedback during skill execution instead of / in addition to using alpha ERD as the cue to initiate movement. Future neurofeedback protocols that target during movement control could employ wearable technologies to assess intervention effects on the execution of regular activities of daily living^40^. They could further record psychological factors related to mood, motivation and self-efficacy to evaluate their role as possible mediating factors of neurofeedback training benefits^18^.

We encourage future efforts to use the CRED-nf checklist^26^ to ensure best practice guidelines are followed and rigorous experimental designs continue to be adopted. We also encourage researchers to continue to pre-register their trials and we hope that the documentation of this experiment will inform more detailed pre-registered study protocols in the future^41,42,43^. Lastly, while the present study focused on motor rehabilitation in PD, the shown proof-of-concept for a multi-session supervised neurofeedback experiment in a home-setting may encourage exploration of other clinical use cases. For instance, home-based neurofeedback training can target cognitive symptoms in neurodegenerative disorders^44^ or mental diseases^45,46^ or aim to facilitate movement in other rehabilitation contexts (e.g., recovery from stroke)^47^. Recent technical advances in mobile EEG^48^ and optical neuroimaging methods such as functional near-infrared spectroscopy (fNIRS)^49,50^ further expand the exciting prospects of mobile, ambulatory, home-based and even dyadic^51^ neurofeedback applications.

### Conclusion

This study provided the first examination of home-based EEG neurofeedback as a potential non-pharmacological, personalised intervention for people with PD. The results provide novel evidence to endorse the feasibility of home-based EEG neurofeedback training to help people with PD to self-regulate preparatory movement-related brain rhythms and serve as a scalable non-invasive, neuromodulatory intervention. Data further indicate that alpha ERD neurofeedback may expedite the initiation and enhance the consistency of fine movements, but our results fall short of providing evidence for transfer benefits to online control of gross movements or wider symptomology reduction. Given the high acceptability and the clear feasibility of home-based neurofeedback in early-stage PD, we see considerable promise for neurofeedback as a non-pharmacological treatment. Larger-scale investigations of neurofeedback in people with PD are warranted and we believe that this founding work can provide a significant blueprint for those future research endeavours.

## Method

### Protocol Registration

The methodology, hypotheses, sampling plan, primary as well as secondary outcomes were prospectively registered on 29^th^ August 2017 (https://www.isrctn.com/ISRCTN16783092).

### Recruitment and sampling plan

Recruitment to the study was initiated by members of the clinical research team who were involved in Movement Disorders Clinics and Parkinson’s Support Groups in the local health board, and who were familiar with study inclusion/exclusion criteria (Table 1) in order to provide initial eligibility assessments. This led to 31 potential participants being identified and consenting to a follow-up conversation with the lead researcher. Four of the potential participants did not respond to the researcher follow-up contact, and four withdrew their interest after contact was made. The remaining 23 potential participants agreed to join a recruitment list to take part in the study. Of these, seven participants were unable to sign-up within the data collection window (e.g., due to illness, personal circumstances or participation in another study); the remaining 16 signed-up and participated in the study. Hence, our sampling plan was based on the number of people with PD we could contact, enrol and who could complete the study within a 7-month data collection period^52^.

### Participants

Sixteen (10 male, 6 female; *M* _age_ = 67.31, *SD* = 9.77 years) participants in the early stage of PD (Hoehn and Yahr stage 1-2; *M* _years since diagnosis_ = 5.06) consented to take part. Fourteen of the participants were receiving treatment with L-Dopa and two participants were receiving other Parkinsonian medication. Importantly, there was no change in medication dosage over the course of the experiment. The experiment received favourable opinions from both the University (reference 16-15660) and the National Health Service Health Research Authority (reference 16/WA/0115) research ethics committees. All participants provided informed consent before taking part. A sensitivity analysis with G*Power 3.1 and its default settings for within-factor effects of a repeated measures analysis of variance (ANOVA)^53^ indicated that a sample size of 13 to 15 (to account for occasional missing data – see statistical analysis section below) would allow detecting within-participant changes of *f*= 0.35 - 0.38 / η_p_^2^ = .62 - .68 (i.e., large size effects^54^) with 80% power for a 3-level repeated measures ANOVA at an alpha level of 0.05. While there are no previous studies of ERD neurofeedback in PD, previous studies of a) the effects of L-Dopa medication on alpha ERD in PD^20^ and b) the effects of alpha ERD neurofeedback in healthy controls^24^, have revealed large effects for within-person changes in cortical activity (our key index of neurofeedback feasibility).

### Design

We employed a fully within-subjects design. Each participant received six home visits from a researcher, with each visit separated by a minimum of 48 hrs. There were two distinct phases of the experiment. The “Test” phase of the experiment comprised home visits 1, 2 and 6. These visits are respectfully labelled as “pre-test A”, “pre-test B” and “post-test”. Participants were on their regular medication during pre-test A (i.e., on-medication) and refrained from taking their medication (overnight withdrawal) ahead of pre-test B (off-medication) and post-test (off-medication). In all visits comprising the Test phase, participants completed assessments of PD symptomatology and performed a force production task (see Precision Motor Task section below) while their cortical activity was measured. The “Training” phase of the experiment comprised home visits 3, 4 and 5. These visits are respectfully labelled as “neurofeedback session 1”, “neurofeedback session 2” and “neurofeedback session 3”. Participants refrained from taking their medication (overnight withdrawal) ahead of all neurofeedback sessions (i.e., off-medication). Each neurofeedback session comprised 12 × 5 min blocks of neurofeedback training, ensuring a total of 1 hr of neurofeedback training per each session. A schematic of the design is provided in Fig 4, and a more detailed description of the neurofeedback session and block factors are provided in the “Neurofeedback Intervention” section below.

**Figure 4.**
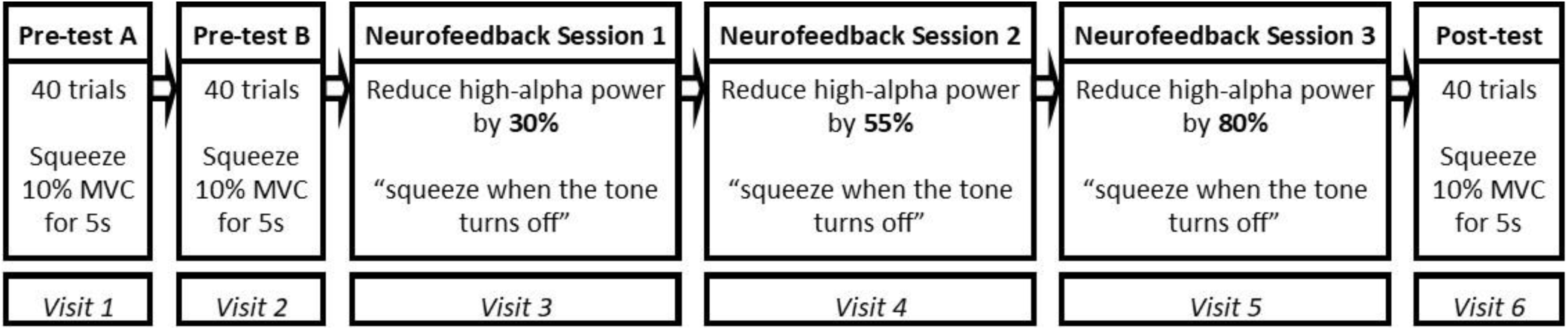
Schematic of experiment design. All participants received six home visits, each separated by at least 48 hrs. The test phase of the experiment comprised visits 1, 2 and 6 and the neurofeedback training phase of the experiment occurred in visits 3, 4 and 5. The target reduction of alpha power was calibrated based on participants’ resting state activity as measured at the beginning of Neurofeedback Session 1.

### Precision Motor Task

During each visit in the Test phase of the experiment, participants performed 40 trials of a precision grip force task. Specifically, they used their dominant hand (right-handed *N* = 14, left-handed *N* = 2) to squeeze a handgrip dynamometer, with the goal of sustaining a force equivalent to 10% of their maximum grip strength (i.e., maximum voluntary contraction; MVC) as accurately as possible for 5 s. Participants commenced each trial by squeezing the dynamometer to trigger a static yellow force line to appear on a computer screen set at 10% of their pre-established MVC. Once the yellow line appeared, participants had to adjust their current grip strength, which was depicted by a dynamic pink line on the screen, to trace the static yellow target line as closely as possible. Both lines remained on the screen for 5 s and then disappeared to indicate the end of the trial. Participants were asked to release their grip at the end of each trial, take a moment to re-focus, and then re-grip the dynamometer to commence the next trial as soon as they felt ready. This precision grip task is commonly used to assess fine motor control^55,56,57^.

### Neurofeedback Intervention

During each visit in the training phase of the experiment, participants received 1 hr of neurofeedback training. At the beginning of the first neurofeedback session, baseline cortical activity was assessed from the C3 electrode site for right-handed participants and the C4 electrode site for left-handed participants. This baseline activity was later used to calibrate the target threshold of the subsequent neurofeedback training visits. Sites C3 and C4 are assumed to approximately correspond to the right- and left-hand motor area, respectively, within the primary motor cortex^27^. We focused our feedback at the C3 (C4 for left-handers) sites because the alpha ERD measured over the primary motor cortex supports movement planning and fine motor control of the hands^17,18^. Additionally, an electrode was placed over the orbicularis oculi muscle of the left eye (right eye for left-handers) to remove eye-blink artefacts, with linked reference electrodes attached to the right and left mastoids and a ground electrode attached to FPz. Recordings were acquired by active electrodes connected to a wireless 4-channel neurofeedback system (Brainquiry PET-4, Nijmegen, The Netherlands). In tandem with cortical recordings, a computer running Bioexplorer software (Cyberevolution, U.K.) used a 6^th^ order Butterworth infinite impulse response 9 to 11 Hz bandpass filter to extract high-alpha power (9-11 Hz) from the EEG signal and fed this back to the participants in the form of an auditory tone^23^. Importantly, the tone was programmed to vary in pitch based on the level of high-alpha power and silence completely when high-alpha power was decreased by 30% (neurofeedback training session 1), 55% (neurofeedback training session 2) and 80% (neurofeedback training session 3), relative to each participant’s individual baseline cortical activity as acquired before the first training session.

These thresholds were based on previous research documenting similar decreases in EEG power during motor preparation^16^, and confirmed via pilot testing which established that they were achievable during our brief intervention. As high-alpha power is inversely related with cortical activity, the progressively more extreme thresholds were designed to encourage increased activation at the C3 (or C4) sites above the primary motor areas, which is characteristic of relatively autonomous and efficient motor preparation^19^. In addition to reducing high-alpha power by the aforementioned thresholds, the system also required <10 µV of 50 Hz activity in the signal (i.e., low mains noise) and the absence of eye-blinks, as detected by the electrode paced adjacent to the eye contralateral to the dominant hand (eye-blinks were detected as >75 µV of 1-7 Hz activity at the eye-electrode), for the tone to silence. These control features helped ensure the signal was being regulated by cognitive processes and was not contaminated by electrical, muscular or eye-blink artefacts^24^.

The auditory neurofeedback training was delivered to participants over 12 × 5-min blocks, each separated by a 2- min break. Participants were seated in a comfortable chair in their home. Each time the thresholds described above were met, the auditory tone was set to silence for 1.5 s and participants were instructed to squeeze the handgrip dynamometer with their dominant hand to produce a grip force equivalent to 10% MVC for 5 s (i.e., to initiate a trial of the precision motor task described in the section above). This instruction was designed to help participants associate the relative increase in cortical activation with the onset of movement.

### Measures

#### Cortical Activity

EEG activity was recorded from the Fz, Cz, C3 and C4 sites on the scalp^58^ during the test phase of the experiment, and from C3 (for right-handers) or C4 (for left-handers) during the neurofeedback training. Recordings were obtained via active electrodes connected to a DC amplifier (Brainquiry PET-4, The Netherlands), with linked reference electrodes attached to the right and left mastoids and a ground electrode attached to FPz. Recording sites were cleaned, abraded, and conductive gel (Signa gel, Parker, Biosense Medical, U.K.) was applied to ensure that electrode impedances were below 10kΩ. The signals were digitized at 24-bit resolution (Brainquiry, The Netherlands) and transmitted via Bluetooth at a sampling rate of 200 Hz to a computer running Bioexplorer (Cyberevolution, U.K.) software. We employed Butterworth infinite impulse response (6th order) bandpass filter at 9-11 Hz (high-alpha power) to extract EEG data from each recording.

#### Precision Handgrip Performance

Performance on the precision motor task was assessed in four ways. We recorded the time in milliseconds (ms) from the onset of each trial to the moment that the force produced by the participant arrived and remained between 9% and 11% of their MVC for 200 ms (i.e., time taken to get into the region of the 10% target – Fig. 5). We interpret this time-based measure as an index of the participants’ ability to plan the movement (e.g., ^30,31^), with shorter times indicating more accurate pre-planning of force and, thus, better performance. We also recorded three error scores. Absolute error was the discrepancy (in % of MVC) between the target force and the produced force; this reflects accuracy (i.e., proximity to target). Constant error was similar, but with the sign of the error (negative for errors below target force and positive for errors above target force) factored in; this reflects bias (i.e., any tendency to undersqueeze or oversqueeze). Finally, variable error was the within-person standard deviation of constant error scores across trials; this reflects consistency (i.e., consistency of bias across trials). In all cases higher scores represent worse performance (i.e., greater error, greater bias, inconsistency) and scores of zero (no error, no bias, perfect consistency) are optimal. These measures were averaged during each trial, with the averaging commencing at the point at which participants’ force arrived and remained between 9% and 11% of their MVC for 200ms and terminating at the end of each 5 s trial (Fig. 5). These error measures thereby reflect the online component of movement control^30,31^. All performance measures were calculated for each trial and then averaged over the total number of trials per visit. This yielded a single score for each measure for each of the six experimental visits.

**Figure 5.**
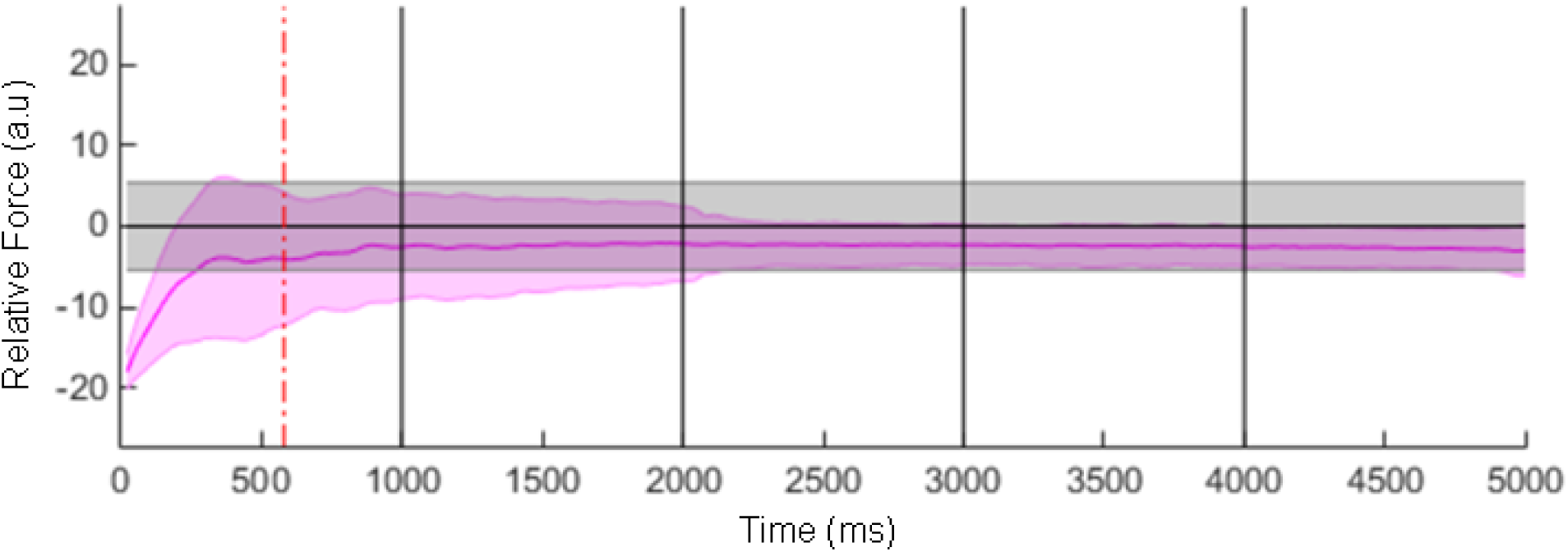
Example force recording from our handgrip task. Solid pink line indicates force produced. Horizontal black line indicates target force (i.e., 10% MVC). Shaded grey area indicates ± 1% of target (i.e., 9-11% MVC). Our time-based measure of movement planning was calculated as the time from the onset of each trial to the moment that the force produced arrived and remained between 9-11% MVC for 200 ms, indicated by the dotted red line on this trace. Movement accuracy (absolute error) and bias (constant error) concerned the discrepancy between the produced force (pink line) and the target force (horizontal black line) while consistency (variable error) concerned the consistency of the produced force to target force discrepancies across trials. All error scores were computed from the dotted red line until the end of each 5 s trial.

#### Motor Symptoms of PD: Self-reported

Participants’ self-reported PD motor symptoms were measured using the Motor Aspects of Experiences of Daily Living (MEDL) questionnaire, which is Part II of the Movement Disorder Society Unified Parkinson’s Disease Rating Scale (MDS-UPDRS)^59^. This subscale comprises 13 items enquiring about difficulties experienced during motor aspects of daily life (e.g., dressing; handwriting; turning in bed) during the past week, with items rated on a 5-point scale ranging from 0 (no problems) to 4 (severe problems). Responses to the 13 items are summed to yield a score ranging from 0 to 52, with higher scores indicating more severe symptomology.

#### Motor Symptoms of PD: Observer rated

We also conducted an objective assessment of motor function using the Motor Examination that forms Part III of the MDS-UPDRS^59^. The experimenter, a trained MDS-UPDRS rater, observed participants as they engaged in 33 standardised tests (e.g., moving each arm from an outstretched position to the tip of the nose; rising from a chair; extending each arm and turning the hands from palm up to palm down), and rated the movements on a 5-point scale ranging from 0 (no problems) to 4 (severe problems). Scores for the 33 tests are summed to yield a total ranging from 0 to 132, with higher scores indicating more severe symptomology.

#### Quality of Life

We assessed self-reported quality of life using the Parkinson’s Disease Quality of Life Questionnaire (PDQ-8)^60^. Participants responded to eight items concerning how often during the past month they experienced certain feelings or symptoms (e.g., had difficulty getting around in public; felt embarrassed in public due to having Parkinson’s disease) on a 5-point scale (0 = *Never,* 1 = *Occasionally,* 2 = *Sometimes,* 3 = *Often,* 4 = *Always*). Scores were summed, divided by the maximum possible score (32), and multiplied by 100 to yield a standardized score ranging between 0 and 100, where higher scores indicate lower quality of life.

#### Participant Acceptability

At the end of the final neurofeedback training session (neurofeedback session 3) we asked participants to verbally respond to the questions: a) “do you think the neurofeedback training has helped you to control your movements?”; b) “would you recommend this type of training to other people with PD?” and c) did you find any strategies to help you control the neurofeedback tone, and if so, what were they? and we recorded participants’ responses with a Dictaphone. These questions were designed to provide some brief qualitative insight into the experience of neurofeedback training among people with PD.

### Procedure

After identifying potential participants (see Recruitment section above) a member of the research team visited each of the 16 individuals who ultimately became participants and they were fully briefed about the experiment before they provided informed consent, and home visits to conduct the experiment were arranged. Participants’ general practitioners were also informed about the study participation and invited to contact the lead consultant on the research team if they had any questions or concerns; no such contacts were made.

Once consent was established, the researcher made six 2-hr visits to the home of each participant in order for them to complete both the test phase (visits 1, 2 and 6) and the neurofeedback training phase (visits 3, 4 and 5) of the experiment, with each visit separated by a minimum of 48 hrs (*M* inter-session-interval = 8.72 ± 3.84 days). The three visits comprising the test phase followed a common general procedure. First, participants were briefed, seated, and fitted with the EEG recording system. We prepared the skin by lightly abrading over the mastoids with exfoliating paste, and with a blunt needle at the scalp sites (Fz, Cz, C3, C4). The sites were then cleaned with an alcohol wipe, conductive gel was applied, and disposable spot electrodes (BlueSensor, Ambu, Denmark) were placed and secured using tape and a lycra cap. The EEG amplifier was attached by an elastic and Velcro strap to the participant’s non-dominant arm. After instrumentation, we assessed the participant’s MVC. Participants were given a handgrip dynamometer (MLT004/ST, AD Instruments, Australia) and they were asked to squeeze the dynamometer with their dominant hand as hard as they could, for a period of 3 s. This process was repeated five times, with a 1-min break to allow recovery between each maximal attempt. We recorded the biggest force generated during the five attempts as the MVC and used that force to calculate the 10% MVC target for the subsequent precision handgrip task. The researcher entered the target threshold into a bespoke computer script designed to control the experiment (Visual Basic, Microsoft, U.S.A.) and after a short break, the participant was instructed about the precision handgrip task. Specifically, they were told that they would see a pink line on the screen, reflecting their grip force, and a static yellow target line, reflecting the target grip force, which was set at 10% of their MVC. They were further told that each time they squeezed the dynamometer, a trial would start, and their goal was to produce a force that ensured their pink line matched the yellow line as quickly as possible, and remained on the yellow line as closely as possible, for 5 s. At the end of each 5 s trial the screen would go blank, and they were to release their grip, re-focus, and commence the next trial whenever they felt ready. Their EEG was recorded continuously from trial 1 to trial 40, and participants were asked to remain still and quiet where possible for the duration of the 40 trials to help optimize the EEG recordings. Participants required an average of 7 mins 42 sec (*SD* = 1 min 8 s) to complete the 40 trials of the handgrip task. After completing the handgrip task, the EEG hardware was removed, participants were asked to complete the MEDL and the PDQ-8 questionnaires, and the researcher took participants through the MDS-UPDRS Part III motor examination.

The three visits comprising the neurofeedback phase of the experiment adopted a general procedure as follows. First, participants were briefed, seated, and fitted with the neurofeedback system. Recording sites were prepared as described previously and electrodes were affixed at the locations described in the Neurofeedback Intervention section above. Next, we assessed the participant’s baseline high-alpha power by asking them to fixate on a cross displayed on a computer screen at eye level, for a period of 5 s. During this time, high-alpha power was monitored. This process was repeated five times and the average was used as their baseline high-alpha power value to calibrate subsequent neurofeedback training sessions. Baseline data was only applied in neurofeedback session one to ensure a comparability across training sessions. Having established individual baselines, the experimenter manually set the threshold for silencing the neurofeedback tone in the neurofeedback software. Participants were then instructed that in the next phase of the experiment they would hear a tone that was contingent on their brainwaves, and which would silence when they produced the type of brainwaves that we theorized would aid their movement. No explicit strategy was provided and participants were asked to try to identify successful regulation strategies (i.e., those that were followed by silenced tone). Further, participants were asked to squeeze the handgrip dynamometer at 10% MVC for 5 s when the tone was silenced in order to associate reduced high-alpha power with the onset of movement. Participants completed the 12 × 5-min blocks of neurofeedback training, with brief comfort breaks as required between each block. The neurofeedback hardware was removed at the end of the final neurofeedback block, and at the end of the final neurofeedback training session, participants were asked to verbally respond to the participant acceptability questions and we recorded their responses on a Dictaphone. At the end of the final home visit participants were thanked for a final time and invited to contact the experimenter for the results of the experiment at the end of the data collection period.

### Statistical Analyses

One participant withdrew from the experiment after pre-test B, and two participants had noisy EEG data for a portion of at least one of the test sessions; these EEG data were removed from the sample prior to analyses. This yielded a sample of 13 participants for EEG analyses during the test-phase, and 15 participants for EEG analyses during the training phase, and for all the remaining behavioural and self-report assessments.

#### Quantitative Analyses

We analysed our quantitative data during the test phase (EEG, precision handgrip performance, PD symptomology), the training phase (EEG, precision handgrip performance), and across the combination of phases to capture all six visits (EEG, precision handgrip performance). In the test phase we had a-priori expectations of quadratic effects (i.e., similar outcomes in pre-test A and post-test, and discrepant outcomes in pre-test B). Accordingly, we ran 3 Test (pre-test A, pre-test B, post-test) repeated measures Polynomial Trend ANOVAs with a focus on the quadratic outcomes. In the training phase we expected linear effects (e.g., progression from one training session to the next). We therefore ran 3 Session (Neurofeedback Session 1-3) Polynomial Trend ANOVAs for our measures of handgrip performance and a 3 Session × 12 Block (EEG during each 5-min block of training) Polynomial Trend ANOVA for cortical activity, with a focus on the linear outcomes. Although the training and test phases reflected distinct components of our experiment with subtly different procedures and predictions, for completeness, we also ran six-level repeated measures ANOVAs (i.e., pre-test A, pre-test B, neurofeedback session 1-3, post-test) for the EEG and performance measures. As we had no firm a-priori predictions about these six-level analyses, we report the univariate ANOVA outcomes, with the Greenhouse-Geisser correction procedure applied wherever the sphericity of variance assumption was violated. Finally, we computed the number of times the neurofeedback tone was silenced in each 5-min training block and subjected these data to a 3 (Session) × 12 (Block) Polynomial Trend ANOVA. This measure reflects the real-time index of neurofeedback as provided to participants^26^ with higher scores indicating better control of brainwaves. For brevity, the results of the tone silence analyses are reported in the supplementary online material.

#### Qualitative Analyses – Participant Acceptability

To assess participant acceptability, we transcribed participants’ responses to the three verbal questions posed at the end of the final neurofeedback training session and we used descriptive and thematic content analyses to summarise the participant views.

## Supporting information

CRED-NF checklist

## Data Availability

The datasets analysed during the current study are available from the corresponding author on reasonable request.

## Code Availability

The Bioexplorer scripts used for the neurofeedback intervention are available from the corresponding author on reasonable request.

## Authors contributions

Conceptualization: [AC, JH, AWP, CAM, PMF, MB, DEJL, DMAM], Methodology: [AC, JH, CL, AWP, CAM, PMF, MB, DEJL, DMAM], Formal analysis and investigation: [AC, CL, EB, DMAM], Writing - original draft preparation: [AC, JH, AWP, DMAM]; Writing - review and editing: [AC, JH, CL, EB, CAM, MB, DEJL, DMAM], Funding acquisition: [AC, JH, AWP, CAM, PMF, MB, DEJL, DMAM], Resources: [AC, JH, CL, EB, AWP, CAM, PMF, MB, DEJL, DMAM], Supervision: [AC, JH, SJ].

## Ethical Approval

Approval was obtained from the National Health Service Wales Research Ethics Committee 4 (Ref 16/WA/0115) and from Bangor University School of Psychology Human Research Ethics Committee. The experiment was performed in accordance with the Declaration of Helsinki ethical standards.

## Consent to participate

Informed consent was obtained from all individual participants included in the study.

## Consent for publication

Participants have consented to the submission of this experiment to scientific journals.

## Availability of data and material

Data and materials are available upon reasonable request to the corresponding author.

## Trial registration

ISRCTN16783092 prospectively registered on 29/08/2017

