## Supplementary Material for "Effects of Home-Based EEG Neurofeedback Training as a Non-Pharmacological Intervention for Parkinson’s Disease"

### Training Phase – Number of Times Neurofeedback Tone Silenced

To test whether participants gained progressively more control over their brainwaves as their neurofeedback training sessions progressed, we analysed the number of times the criteria for silencing the neurofeedback tone were met using a 3 (neurofeedback session) × 12 (block) Polynomial Trend ANOVA. Results revealed a significant linear trend for Session, $F(1,12) = 24.53$, $p = .000$, $\eta_p^2 = .67$, showing a slight decrease in the number of times the tone was silenced per block from the first to the third neurofeedback training session ($M_{\text{session 1}} = 28.50$, $M_{\text{session 2}} = 26.41$, $M_{\text{session 3}} = 24.92$ per block). This evidences that the shaping of the neurofeedback thresholds across session (i.e., participants had to produce a stronger decrease in high-alpha power to silence the tone in session 2, and again in session 3, compared to their preceding session) successfully increased the challenge of the training as sessions progressed, while maintaining a level where participants were able to silence the tone frequently during each 5 min block (approximately every 12 s). Most importantly, there was a significant linear trend for Block, $F(1,12) = 5.65$, $p = .03$, $\eta_p^2 = .32$ characterized by an increase in the number of times the tone was silenced per block from the start to the end of the training sessions (Supplementary Fig. 1). This provides evidence that participants became more successful at producing the prescribed brainwaves over the course of each training session. The interaction effect was not significant, indicating that the linear effect of block was similar across all three neurofeedback sessions.

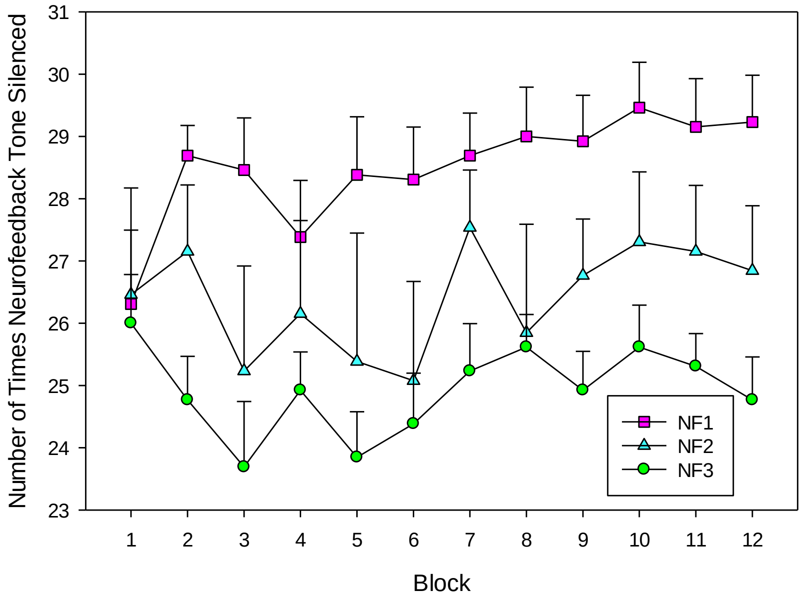


*Supplementary Figure 1.* Mean number of times the neurofeedback tone was silenced per each three-minute block of neurofeedback training across all training sessions. Error bars indicate standard error of the means.

**Qualitative Analysis**

Thematic content analysis was performed on the responses to the question "did you find any strategies to help you control the neurofeedback tone, and if so, what were they?" Results are summarised in Supplementary Table 1

**Supplementary Table 1**. Thematic content analysis of neurofeedback strategies.

| Theme | Number of Participants | Examples |
|---|---|---|
| Concentration / Focus | 9 out of 15 | "I focused on the line" |
| | | "I tried to sense my brain and focus" |
| | | "I created a focus point" |
| | | "I concentrated on the line and nothing else" |
| | | "I concentrated on the screen" |
| | | "I concentrated on the number 1 on the scale on the screen" |
| Relaxation | 7 out of 15 | "I relaxed my mind" |
| | | "I was relaxing and emptying the mind" |
| Breathing | 3 out of 15 | "I focused on yoga style breathing" |
| | | "I slowed my breathing down" |
| Imagery | 2 out of 15 | "I was thinking about the beach and my grandchildren" |
| | | "I imagined a calm sea" |
| Strategies to prevent artifacts | 2 out of 15 | "I tried to avoid blinking and swallowing" |
| | | "I was focused on not blinking" |
| Self-talk | 1 out of 15 | "I repeated the words *keep quiet* in my mind" |
| Defocusing | 1 out of 15 | "I tried to defocus and blank out" |
| No strategies identified | 3 out of 15 | "I wasn't consistent. I would just get a bit distracted by congratulating myself". |

*Note.* While concentration/focus and relaxation may seem to oppose one another, they often came as a pair, for example, one participant noted that "I concentrated on the number one on the scale on the screen and I used that to help me calm down and block everything else out", while another participant noted, "I was trying to empty my mind, relax, sense my brain, and focus, like mindfulness". This could point towards a gating by inhibition strategy where resources are intensified towards a specific aspect of the environment (e.g., a point on the screen), and aided by simultaneous active inhibition strategies (e.g., mind emptying) to minimize monitoring of any other aspects of the environment.
