## Supplementary material for "Effects of Home-Based EEG Neurofeedback Training as a Non-Pharmacological Intervention for Parkinson’s Disease": CRED-NF checklist

### CRED-nf checklist summary

*02 February, 2024*

**Manuscript title:** Effects of Home-Based EEG Neurofeedback Training as a Non-Pharmacological Treatment for Motor Symptoms of Parkinson's Disease
**Corresponding Author:** Andrew Cooke

| Item No. | Checklist item | Manuscript Details |
|---|---|---|
| **Pre-experiment** | | |
| 1a | Pre-register experimental protocol and planned analyses | The methodology, hypotheses, sampling plan, primary as well as secondary outcomes were registered after funding acquisition and ethical approval but before data collection commenced (https://www.isrctn.com/ISRCTN16783092). |
| 1b | Justify sample size | Hence, our sampling plan was based on the number of patients who we could contact and enrol through support groups within the planned project period. |
| **Control groups** | | |
| 2a | Employ control group(s) or control condition(s) | *This experiment did not include a control group or control condition* |
| 2b | When leveraging experimental designs where a double-blind is possible, use a double-blind | *NA: A double-blind was not appropriate for this experiment* |
| 2c | Blind those who rate the outcomes | *NA: There was only one participant group* |
| | Blind those who analyse the data | *NA: There was only one participant group* |
| 2d | Examine to what extent participants and experimenters remain blinded | *NA: There was only one participant group* |
| 2e | In clinical efficacy studies, employ a standard-of-care intervention group as a benchmark for improvement | *NA: This is not a clinical efficacy study* |
| **Control measures** | | |
| 3a | Collect data on psychosocial factors | *Psychosocial factors were not measured* |

| | | |
|---|---|---|
| 3b | Report whether participants were provided with a strategy | Participants were instructed that in the next phase of the experiment they would hear a tone that was contingent on their brainwaves, and which would silence when they produced the type of brainwaves that we theorized would aid their movement. No explicit strategy was provided. We asked participants were asked to try to identify successful regulation strategies, i.e., those that were followed by silenced tone. |
| 3c | Report the strategies participants used | Participants were then instructed that in the next phase of the experiment they would hear a tone that was contingent on their brainwaves, and which would silence when they produced the type of brainwaves that we theorized would aid their movement. No explicit strategy was provided and participants were asked to try to identify successful regulation strategies (i.e., those that were followed by silenced tone). |
| 3d | Report methods used for online-data processing and artifact correction | An electrode was placed over the orbicularis oculi muscle of the left eye (right eye for left-handers) to remove eyeblink artefacts, with linked reference electrodes attached to the right and left mastoids and a ground electrode attached to FPz. We focused our feedback at the C3 (C4 for left-handers) sites because the alpha ERD measured over the primary motor cortex supports movement planning and fine motor control of the hands (Defebvre et al., 1994; Mehler, 2022). Recordings were acquired by active electrodes connected to a wireless 4-channel neurofeedback system (Brainquiry PET-4, Nijmegen, The Netherlands). ... In addition to reducing high-alpha power by the aforementioned thresholds, the system also required <10 V of 50Hz activity in the signal (i.e., low impedance) and the absence of eye-blinks, as detected by the electrode paced adjacent to the eye contralateral to the dominant hand (eye-blinks were detected as >75 V of 1-7 Hz activity at the eye-electrode), for the tone to silence. These control features helped ensure the signal was being regulated by cognitive processes and was not contaminated by electrical, muscular or eye-blink artefacts (Ring et al., 2015). |
| 3e | Report condition and group effects for artifacts | Condition and group effects for artifacts were not measured, or not reported in the manuscript. |
| **Feedback specifications** | | |
| 4a | Report how the online-feature extraction was defined | In tandem with cortical recordings, a computer running Bioexplorer software (Cyberevolution, U.K.) used a 6th order Butterworth infinite impulse response 9-11 Hz bandpass filter to extract high-alpha power (9-11 Hz) from the EEG signal and fed this back to the participants in the form of an auditory tone (Ring et al. 2015). |
| 4b | Report and justify the reinforcement schedule | Importantly, the tone was programmed to vary in pitch based on the level of high-alpha power and silence completely when high-alpha power was decreased by 30% (neurofeedback training session 1), 55% (neurofeedback training session 2) and 80% (neurofeedback training session 3), relative to each participant's individual baseline cortical activity as acquired before the first training session. |

| | | |
|---|---|---|
| 4c | Report the feedback modality and content | See above |
| 4d | Collect and report all brain activity variable(s) and/or contrasts used for feedback, as displayed to experimental participants | See above |
| 4e | Report the hardware and software used | Recordings were acquired by active electrodes connected to a wireless 4-channel neurofeedback system (Brainquiry PET-4, Nijmegen, The Netherlands). |
| 5a | Report neurofeedback regulation success based on the feedback signal | SUPPLEMENTARY MATERIAL Training Phase – Number of Times Neurofeedback Tone Silenced To test whether participants gained progressively more control over their brainwaves as their neurofeedback training sessions progressed, we analysed the number of times the criteria for silencing the neurofeedback tone were met using a 3 (neurofeedback session) × 12 (block) Polynomial Trend ANOVA. Results revealed a significant linear trend for Session, $F(1,12) = 24.53$, $p < .001$, p2 = .67, showing a slight decrease in the number of times the tone was silenced per block from the first to the third neurofeedback training session (Msession 1 = 28.50, Msession 2 = 26.41, Msession 3 = 24.92 per block). This evidences that the shaping of the neurofeedback thresholds across session (i.e., participants had to produce a stronger decrease in high-alpha power to silence the tone in session 2, and again in session 3, compared to their preceding session) successfully increased the challenge of the training as sessions progressed, while maintaining a level where participants were able to silence the tone frequently during each 5 min block (approximately every 12 sec). Most importantly, there was a significant linear trend for Block, $F(1,12) = 5.65$, $p < .05$, p2 = .32 characterized by an increase in the number of times the tone was silenced |
| | | per block from the start to the end of the training sessions (Supplementary Fig. 1). |
| 5b | Plot within-session and between-session regulation blocks of feedback variable(s), as well as pre-to-post resting baselines or contrasts | Supplementary Figure 1. Mean number of times the neurofeedback tone was silenced per each three-minute block of neurofeedback training across all training sessions. Error bars indicate standard error of the means. |
| 5c | Statistically compare the experimental condition/group to the control condition(s)/group(s) (not only each group to baseline measures) | *NA: There was only one participant group* |

| | | |
|---|---|---|
| 6a | Include measures of clinical or behavioural significance, defined a priori, and describe whether they were reached | Precision Handgrip Performance Test phase. Repeated measures Polynomial Trend ANOVAs did not reveal the hypothesised quadratic effects for absolute error (F(1,14) = 2.63, p =.13, p2 = .16, constant error (F(1,14) = 0.99, p =.34, p2 = .07), variable error (F(1,14) = 1.23, p =.28, p2 = .08), or movement planning time (F(1,14) = 0.39, p =.55, p2 = .03). There were, however, significant linear effects for variable error (F(1,14) = 5.26, p <.05, p2 = .27) and for movement planning time (F(1,14) = 4.88, p <.05, p2 = .26). Inspection of the means revealed that participants were able to produce grip forces typically within 1% MVC of their target, but with a slight bias to under-squeeze the handgrip dynamometer. Importantly, their movement planning times reduced, and they became more consistent from the pre-tests to the post-test, providing some evidence of improved performance across the Test phase (Figs. 4A, 4D, 4G, 4J). Training Phase. Repeated measures Polynomial Trend ANOVAs did not reveal the hypothesised linear effects for absolute error (F(1,14) = 0.84, p =.37, p2 = .06), constant error (F(1,14) = 1.12, p =.31, p2 = .07) or variable error (F(1,14) = 1.47, p =.25, p2 = .10). There was a significant linear effect for movement planning time (F(1,14) = 14.32, p <.05, p2 = .51). This significant effect w |
| | | as characterized by an improvement in performance (i.e., shorter planning times) from neurofeedback training session 1 to training session 2, and again from training session 2 to training session 3 (Fig. 4K). All visits. Six-level repeated measures ANOVAs to compare performances across all experimental visits revealed no significant effects for absolute error (F(5,70) = 1.87, p =.19, p2 = .12), constant error (F(5,70) = 1.65, p =.22, p2 = .11) or variable error (F(5,70) = 2.77, p =.07, p2 = .16). There was a significant effect for movement planning time (F(5,70) = 4.40, p <.01, p2 = .24). This was characterized by a significant linear trend (F(1,14) = 9.04, p<.01, p2 = .39), with improvements in performance over the course of the experiment, and which were particularly evident during the training phase (Fig. 4L). … MDS-UPDRS and PDQ-8. The mean scores for MDS-UPDRS Part II (motor aspects of experiences of daily living), the MDS-UPDRS Part III (motor examination) and the PDQ-8 are presented in Table 2. Polynomial trend ANOVAs revealed significant quadratic and linear trends for the MDS-UPDRS Part III, (Quadratic: F(1,14) = 5.36, p <.05, p2 = .28; Linear: F(1,14) = 8.28, p <.05, p2 = .37) with the effect sizes indicating that the linear trend was stronger compared to the quadratic trend. However, this |
| | | effect was driven by the least severe symptomatology observed in the (“ON” medication) pre-test A, and most severe symptomology observed in the (“OFF” medication) post-test (Table 2). No significant trends emerged for the self-reported measures, MDS-UPDRS Part II (Quadratic: F(1,14) = 0.71, p =.41, p2 = .05; Linear: F(1,14) = 1.70, p =.21, p2 = .11), PDQ-8, (Quadratic: F(1,14) = 1.09, p =.32, p2 = .07; Linear: F(1,14) = 1.11, p =.31, p2 = .07). |

| | | |
|---|---|---|
| 6b | Run correlational analyses between regulation success and behavioural outcomes | *This manuscript does not compare regulation success and behavioural outcomes* |
| **Data storage** | | |
| 7a | Upload all materials, analysis scripts, code, and raw data used for analyses, as well as final values, to an open access data repository, when feasible | *No additional documents related to the materials, analysis scripts, code, raw data, or final values are available for this manuscript* |
